# Machine Learning–Enabled Smartphone CRISPR-Cas12a Lateral Flow Platform for Sensitive Detection of Circulating HPV DNA

**DOI:** 10.64898/2026.03.17.26348658

**Authors:** Jipei Liao, Jesse Rima, Aarush Sharma, Jen-Hui Tsou, Feng Jiang

## Abstract

Persistent infection with high-risk human papillomavirus (HPV) is the primary cause of cervical cancer and other HPV-related malignancies. Effective screening and early detection of HPV, particularly in point-of-care (POC) settings, can reduce disease progression and associated mortality. Although PCR-based assays provide high sensitivity, their dependence on centralized laboratory infrastructure limits accessibility in POC settings. CRISPR-Cas diagnostics enable programmable, isothermal detection of HPV with lateral flow assay (LFA) readouts; however, visual interpretation of faint bands can be subjective and inconsistent. Our objective was to develop a machine learning (ML)-enhanced, smartphone-native CRISPR-LFA platform for highly sensitive and reliable detection of HPV DNA in plasma. A smartphone-based diagnostic system integrating CRISPR-LFA with a ML framework was developed using standardized image acquisition within a light-controlled enclosure. Radiomics-inspired strip features were extracted and analyzed using a multivariable logistic regression model. A total of 150 plasma samples were used for model development and 60 independent samples for validation. An optimized model was developed that had 96.7% sensitivity and 100% specificity for detection of HPV DNA. The smartphone-enabled CRISPR platform demonstrated higher sensitivity than visual interpretation, particularly for faint-band results, and reduced false positives. Validation in the independent cohort confirmed the robustness of the assay. Performance remained stable across smartphone models, lighting conditions, and operators, and on-device inference enabled reliable operation. In sum, the smartphone-integrated CRISPR-LFA platform can facilitate accurate and reliable detection of plasma HPV DNA in POC settings and has the potential to enhance early detection, prevention, and treatment of cervical cancer.

## Introduction

Cervical cancer arises mainly from persistent infection with oncogenic human papillomavirus (HPV), especially genotypes 16 and 18, which are also implicated in multiple anogenital and oropharyngeal malignancies ^1, 2^. Although vaccination programs and cytology-based screening have reduced cervical cancer incidence in well-resourced regions, prevention efforts remain inadequate in many low-resource and underserved settings. Early and accurate detection of high-risk HPV infection is essential for effective prevention, timely intervention, and reducing disease progression and mortality. However, current HPV detection strategies, such as polymerase chain reaction (PCR), require centralized laboratory infrastructure and specialized personnel, limiting their applicability in point-of-care (POC) settings.

Clustered Regularly Interspaced Short Palindromic Repeats (CRISPR), an adaptive immune system originally discovered in bacteria and archaea, has been harnessed for nucleic acid detection^3^. CRISPR-based molecular diagnostics provide programmable target recognition, compatibility with isothermal amplification, and lateral flow assay (LFA) readout capability, emerging as robust alternatives to PCR for rapid detection of DNA ^4^. We have demonstrated that CRISPR-Cas detection platforms achieve PCR-comparable analytical sensitivity while significantly reducing workflow complexity for the detection of viral and DNA targets, including HPV DNA ^5-10^. However, a key translational barrier of CRISPR-based diagnostics is that lateral flow strip interpretation remains visually subjective and operator-dependent. Faint or borderline bands in low-abundance nucleic acid assays may introduce inter-reader variability, reduce reproducibility, and increase false negatives. This limitation is especially significant in cervical cancer screening applications, where subtle signal differences may have important clinical implications in POC settings.

Recent efforts have applied deep learning models to automate interpretation of CRISPR-LFA images ^11-13^. While demonstrating promising classification accuracy, the approaches often employ complex convolutional architectures with limited interpretability, which may present challenges for assay optimization, reproducibility, and regulatory validation^13^. Interestingly, radiomics is a quantitative imaging approach that extracts high-dimensional, reproducible, and interpretable features from medical images^14^. It has transformed image analysis by converting imaging data into mineable features and integrating them into statistically calibrated predictive models ^14-16^. Using a radiomics-based framework, we evaluated feature stability, removed highly correlated variables, and performed dimensionality reduction of features derived from CT images ^17^. We then developed a reproducible and clinically translatable radiomic model to classify pulmonary nodules on low-dose CT and improve early lung cancer detection ^17^. Building on our previous efforts^5-10, 17^, the present study aimed to develop a smartphone-based CRISPR-LFA assay for plasma HPV detection, incorporating radiomics and an interpretable machine learning (ML) framework. The developed assay shows the potential to improve CRISPR-LFA testing by enabling objective and quantitative detection of HPV DNA with enhanced sensitivity and reliability.

## Materials and Methods

### Study Overview

The overall design comprised four integrated components (Fig. 1). First, a CRISPR-LFA workflow was used for plasma-based HPV detection. Second, a standardized smartphone imaging system was implemented to ensure reproducible optical acquisition. Third, predefined quantitative strip features were extracted and analyzed using an on-device ML model to generate calibrated probabilistic predictions. Finally, encrypted data storage and secure transmission capabilities were incorporated to enable decentralized testing and centralized epidemiologic oversight.

**Figure 1.**
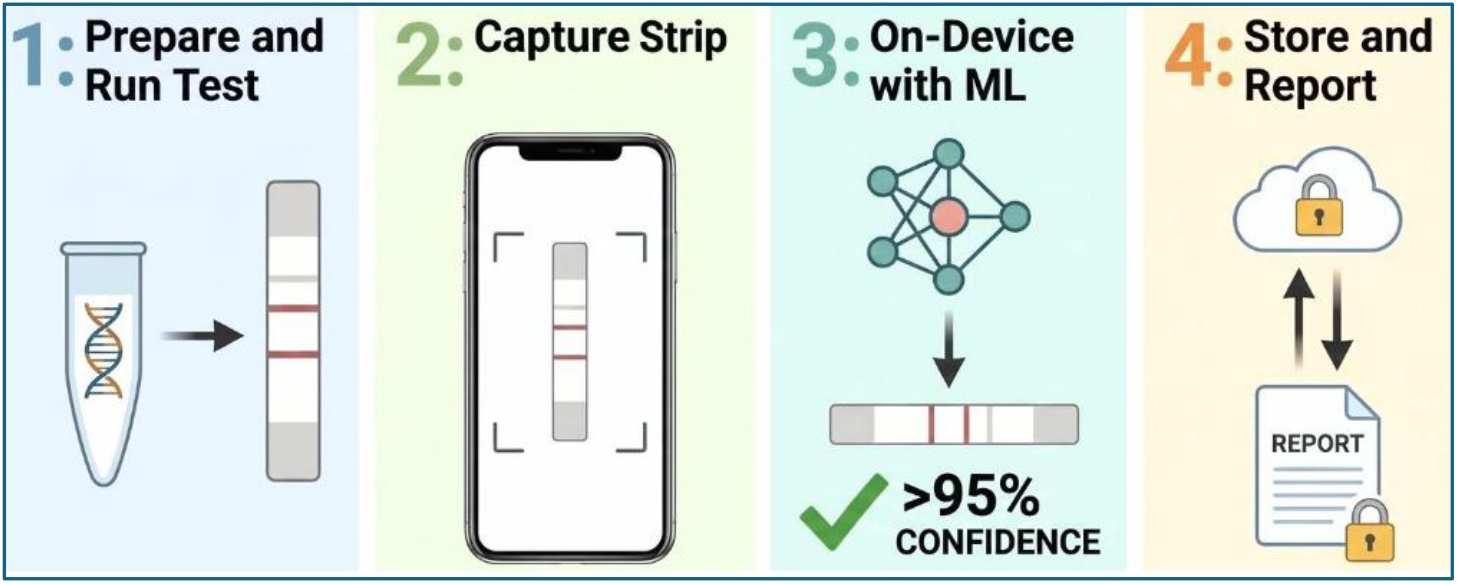
Schematic overview of the integrated smartphone-enabled CRISPR-ML workflow for sensitive detection of circulating HPV DNA. (1) CRISPR-LFA with strip visualization. (2) Standardized smartphone imaging using a light-controlled enclosure. (3) On-device ML analysis of predefined strip regions to generate calibrated probabilistic HPV predictions. (4) Encrypted data storage and secure transmission to support structured reporting, surveillance, and scalable field deployment.

### Human Subjects and Clinical Specimens

The study protocol (22-8356) was reviewed and approved by the Advarra Institutional Review Board (Advarra, Columbia, MD). Peripheral blood was collected in Ethylenediaminetetraacetic acid (EDTA) tubes (Fisher Scientific. Pittsburgh, PA), and plasma was isolated by centrifugation and stored at −80 °C until analysis. A total of 150 plasma samples were used for model development (75 HPV-positive and 75 HPV-negative), alongside an independent validation cohort of 60 samples (30 HPV-positive and 30 HPV-negative) (Supplementary Table S1). Among the 75 HPV-positive development cases, 60 were HPV 16-positive and 15 were HPV 18-positive. Of the 30 validation cases, 23 were HPV 16-positive and seven were HPV 18-positive. The mean age was 39.25 ± 7.39 and 38.25 ± 6.15 years for HPV-positive and HPV-negative subjects, respectively, in the development cohort, and 38.75 ± 9.33 and 39.46 ± 9.28 years in the validation cohort. HPV-positive cases presented with squamous cell carcinoma and were staged according to standard clinical guidelines. Stage distribution in the development cohort included stage 0 (n = 14), A1 (n = 13), IA2 (n = 12), II2 (n = 15), III (n = 11), and IV (n = 10). Corresponding counts in the validation cohort were 5, 6, 4, 7, 5, and 3, respectively.

### CRISPR-LFA Assay for Circulating HPV DNA

Plasma HPV16 and HPV18 DNA was detected using a CRISPR-Cas12a–based LFA workflow as described in our previous studies ^5-7^. Briefly, plasma samples underwent DNA isolation followed by heat treatment at 95 °C for 5 minutes prior to amplification. Recombinase polymerase amplification (RPA) targeting genomic regions of HPV16 and HPV18 was performed at 37 °C for 20 minutes. The amplified products were then incubated with LbCas12a-crRNA complexes (EnGen® Lba Cas12a, New England Biolabs, Ipswich, MA) in the presence of a FAM-biotin-labeled reporter probe (Integrated DNA Technologies, Coralville, IA), enabling target-dependent collateral cleavage-based signal generation. Following the Cas12a reaction, lateral flow strips (Milenia HybriDetect, TwistDx/Milenia Biotec, Germany) were immersed directly into the reaction mixture and allowed to develop for 1-5 minutes at room temperature. Formation of test and control lines was initially confirmed by visual inspection prior to standardized digital image acquisition for quantitative analysis.

### Standardized Smartphone-Based Image Acquisition

Images were acquired using a custom-designed, light-controlled enclosure to standardize strip positioning and illumination (Supplementary Figure 1). The enclosure featured a fixed smartphone mount to maintain consistent camera-to-strip distance and alignment, integrated daylight-balanced LED lighting (5000-6500 K) to minimize ambient variability, and a dedicated slot for strip placement. Fabricated from 3D-printed polylactic acid (PLA) (Hatchbox 3D, Los Angeles, CA) with a matte black interior to reduce reflection and optical scattering, the enclosure was 18 cm x 10 cm x 8 cm in size and was powered by a rechargeable lithium battery module. The smartphone mount and strip holder were fixed within the enclosure to maintain consistent geometry and working distance across acquisitions. The estimated material cost per unit was approximately $22 USD. Images were acquired using iPhone devices (Apple Inc., Cupertino, CA, USA; multiple models tested) mounted in the fixed position. Camera settings were standardized, including locked exposure, fixed white balance, disabled HDR (High Dynamic Range), and fixed focus. A custom iOS application written in Swift was used to capture images and process data.

### Image Processing and Feature Extraction

Images were converted to grayscale and spatially aligned using fixed-geometry cropping. Predefined regions of interest (ROIs) corresponding to the test band, control band, and adjacent background were extracted. Quantitative strip features were computed using a radiomics-inspired, interpretable feature framework as described in our previous report ^17^. Feature selection and model reduction were conducted using a framework incorporating stability assessment, correlation filtering, and parsimonious model construction ^17^. All algorithms were implemented using iOS libraries to ensure deterministic, device-consistent computation.

### ML Model Development

To reduce dimensionality and mitigate overfitting, highly correlated features (Pearson r ≥ 0.85) were identified and removed using hierarchical clustering ^17^. The remaining candidate features were then subjected to least absolute shrinkage and selection operator (LASSO) regression within repeated cross-validation to identify a parsimonious and stable predictor subset. A multivariable logistic regression classifier was subsequently trained using the selected features to generate probabilistic predictions of HPV positivity. Model development was conducted in Python using the scikit-learn framework, employing repeated 10-fold cross-validation (10 repetitions) to ensure stability and reliable internal validation. Model calibration was rigorously evaluated to assess agreement between predicted probabilities and observed outcomes. Calibration performance was examined using calibration plots, estimation of calibration slope and interception, calculation of the Brier score, and the Hosmer-Lemeshow goodness-of-fit test. To quantify potential optimism and adjust performance estimates accordingly, bootstrap resampling with 10,000 iterations was performed to generate optimism-corrected discrimination and calibration metrics.

### On-Device ML Inference Implementation

Following model development, the finalized logistic regression coefficients were embedded within the smartphone application to enable fully self-contained inference. On-device prediction was implemented using the logistic function: 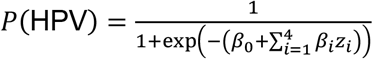 where *z*_*i*_represents standardized quantitative strip features and *β*_*i*_denotes fixed model coefficients derived from the training dataset. Continuous predictors were standardized using development cohort parameters: 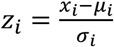 The finalized regression coefficients were derived from the multivariable logistic regression model trained in the development cohort using repeated cross-validation. Because the predictors were standardized before model fitting, the reported coefficients reflect standardized feature values. The standardization parameters are provided to allow full reproducibility of the analysis. All coefficients and preprocessing parameters were embedded within the application to maintain numerical consistency between on-device inference and the validated Python reference implementation. All image processing, feature extraction, and probabilistic inference were performed locally without reliance on proprietary or third-party artificial intelligence (AI) software development kits (SDKs), cloud-based computation, or external servers.

### Data Storage and Secure Transmission

Each completed test generated a structured digital record containing the predicted probability score, binary classification result, timestamp, device identifier, and optional geolocation metadata. These data elements were encrypted and securely stored locally using native iOS data protection frameworks to ensure confidentiality and integrity. When network connectivity was available, encrypted test records were transmitted via secure HTTPS (HyperText Transfer Protocol Secure) protocols to a centralized surveillance database for aggregation and epidemiological monitoring. In the absence of network access, results were cached locally and automatically synchronized upon reconnection, preserving data continuity and minimizing loss during intermittent connectivity. This architecture supports secure and structured data capture and facilitates integration into decentralized public health surveillance systems.

### Validation Strategy

Model performance was evaluated using internal and external validation procedures. Discriminative ability was assessed by receiver operating characteristic (ROC) analysis, with area under the curve (AUC) estimates and 95% confidence intervals calculated using the DeLong method. Sensitivity and specificity were reported with Wilson score confidence intervals to ensure accurate estimation of binomial proportions. To account for potential model optimism, bootstrap resampling with 10,000 iterations was performed. The validation was conducted using an independent cohort to assess generalizability beyond the development dataset. Field robustness testing was used to examine model stability across different smartphone models, lighting conditions, and independent operators to simulate real-world deployment.

### PCR Analysis of HPV DNA in Plasma

Quantification of circulating HPV DNA was determined using a qPCR assay targeting the L1 genotype-specific regions of HPV16 and HPV18, as previously described ^5, 18^. Each PCR reaction was performed in a total volume of 50 µL containing 1× TaqMan Universal PCR Master Mix, genotype-specific primer pairs, and serially diluted HPV plasmid standards using a Bio-Rad CFX96 real-time PCR system (Bio-Rad Laboratories, Hercules, CA, USA). Thermal cycling conditions consisted of an initial denaturation step at 95 °C for 10 minutes followed by 45 amplification cycles of 95 °C, 50 °C, and 72 °C. Cycle threshold (Ct) values were recorded for each reaction, with lower Ct values indicating higher concentrations of HPV DNA in plasma. Samples showing detectable amplification within 39 PCR cycles were classified as HPV-positive, as previously described ^5^. The LOD of this PCR assay was previously determined to be approximately 150 copies/µL (0.24 fM) for HPV16 and 170 copies/µL (0.28 fM) for HPV18 ^5, 18^. PCR results served as the reference standard for defining HPV status in the evaluation of CRISPR-LFA assay performance.

### Statistical Analysis

A total of 150 samples (75 per group) were required to achieve 80% statistical power to demonstrate non-inferiority of sensitivity and specificity relative to a prespecified benchmark of 95%, using a one-sided α of 0.025 and a 5% non-inferiority margin under binomial assumptions. Statistical analyses were performed using Python (v3.11.6; scikit-learn 1.3.2, SciPy 1.11.4, statsmodels 0.14.1) and R (v4.3.2). Continuous variables are reported as mean ± standard deviation (SD), and categorical variables as counts and percentages. Between-group comparisons were conducted using two-sided t-tests or nonparametric equivalents, as appropriate, with p < 0.05 considered statistically significant. Mixed-effects models were used to evaluate the effects of device type, lighting conditions, and operator variability on classification accuracy.

## RESULTS

### CRISPR-LFA Assay for Circulating HPV DNA

In the training cohort comprising 150 plasma samples, 75 were HPV-positive and 75 were HPV-negative controls. Among the 75 HPV-positive cases, 60 were positive for HPV16 and 15 were positive for HPV18, with no samples harboring both genotypes. Visually interpretable CRISPR-LFA test bands were observed in 55 of 60 HPV16-positive samples, whereas five samples displayed faint bands near the threshold of visual detection (Fig. 2A-B). Similarly, 12 of 15 HPV18-positive samples showed clearly interpretable test bands, while three samples exhibited faint bands close to the detection limit (Fig. 2A-B). qPCR analysis of these eight faint-band samples yielded Ct values ranging from 35 to 38, close to the positivity cutoff for HPV16 and HPV18. Based on the previously established calibration curve ^5^, these Ct values correspond to low HPV DNA concentrations of approximately 150 copies/µL, which are near the analytical LOD of the CRISPR-Cas12a assay (approximately 150–170 DNA copies/µL)^5^. Consequently, low viral DNA levels produced weak CRISPR-LFA signals that were difficult to interpret visually, leading to false negatives in these eight faint-band samples. As a result, visually interpreted CRISPR-LFA detected 67 of 75 HPV-positive samples, corresponding to an overall sensitivity of 89.3% (Table 1). No false-positive results were observed among the 75 HPV-negative controls, yielding a specificity of 100% for both HPV16 and HPV18 detection (Fig. 2C)..

**Table 1.** Diagnostic Performance of Visually Interpreted CRISPR-LFA for HPV16 and HPV18 Detection in the Development Cohort ^*^.

| HPV Detection Category | True HPV-16 Positive (n=60) | True HPV-18 Positive (n=15) | True Positive (n=75) | False Positive cases (n=75) | True Negative (n=75) | Sensitivity (%) for HPV-16 | Sensitivity (%) for HPV-18 | Specificity (%) |
| --- | --- | --- | --- | --- | --- | --- | --- | --- |
| HPV16 | 55 | - | 55 | 0 | 75 | 91.7% | - | 100% |
| HPV18 | - | 12 | 12 | 0 | 75 | - | 80.0% | 100% |

\* Overall sensitivity calculated as 89.3% (67/75) for detection of either HPV16 or HPV18 with 100% specificity.

**Figure 2.**
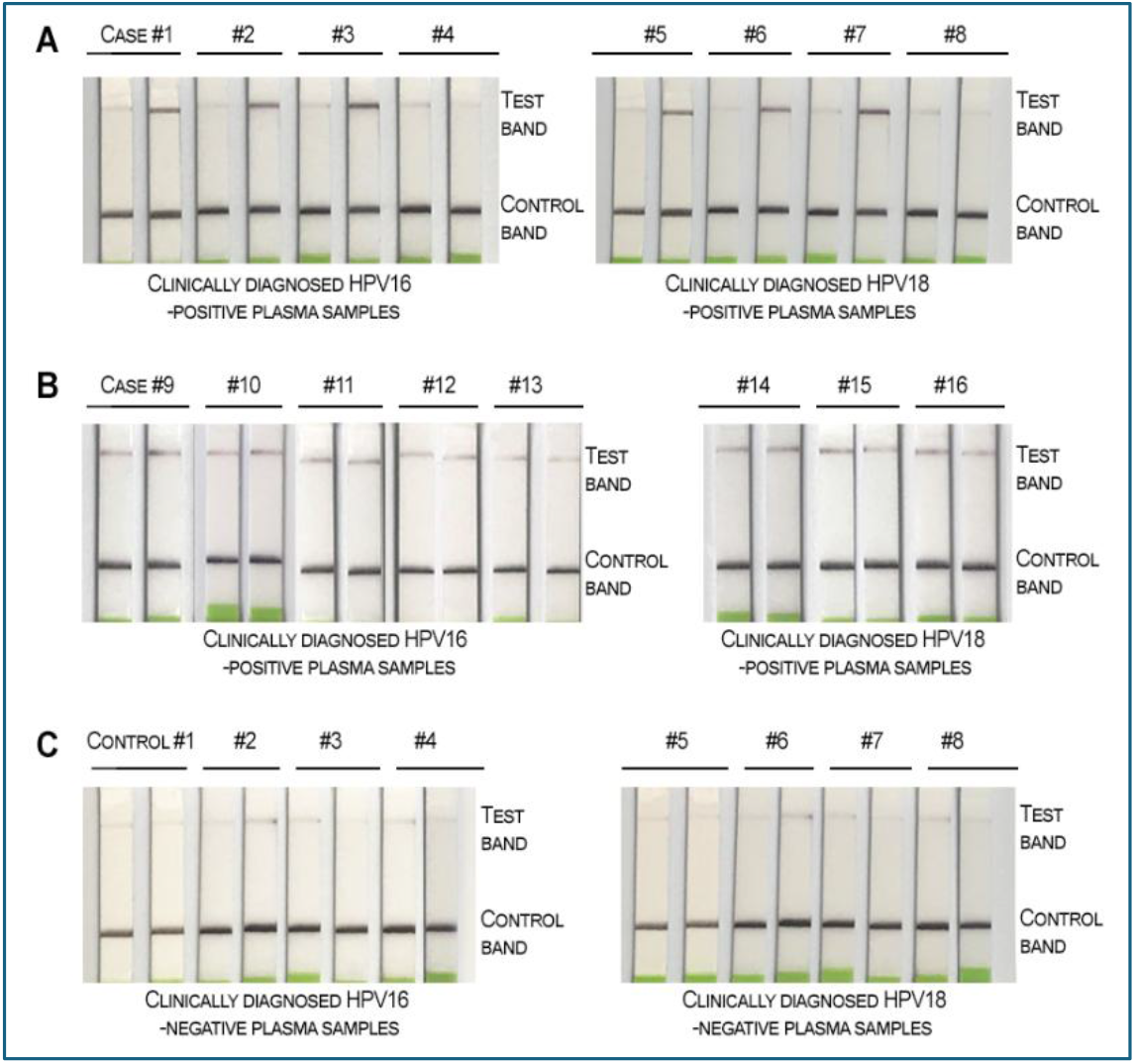
Representative CRISPR-LFA results for plasma HPV detection. Each lateral flow strip contains a test band (upper band) indicating HPV target detection and a control band (lower band) confirming valid assay performance. For each plasma sample, two strips are shown: the left strip represents the control using mutated crRNA, and the right strip represents the target-specific crRNA for HPV16 or HPV18 detection. (A) Clear positive test bands in clinically diagnosed HPV16-positive plasma samples (Cases #1-#4) and HPV18-positive plasma samples (Cases #5-#8). (B) Borderline visual signals or faint test bands were observed in clinically diagnosed HPV16-positive (Cases #9-#13) and HPV18-positive (Cases #14-#16) plasma samples. Near-threshold signal intensity impaired interpretive consistency and contributed to false-negative results. (C) Clinically diagnosed HPV16-negative (left panel) and HPV18-negative (right panel) plasma samples showing absence of test bands.

### Standardized Smartphone-Based Image Acquisition

To ensure reproducible optical conditions, images were acquired using a light-controlled enclosure with fixed cassette positioning and standardized LED illumination. The camera-to-strip distance was maintained at 12.6 ± 0.2 cm across all acquisitions. Illumination uniformity across the strip window demonstrated minimal spatial variability, with a pixel intensity coefficient of variation (CV) <4.1% (Supplementary Figure 2). Repeated imaging of the same strip (n = 20) showed band position deviation <1.5 pixels, confirming geometric stability (Supplementary Figure 2). Internal illumination remained stable at 620 ± 12 lux over 60 minutes of battery-powered operation, indicating temporal consistency. Across three iPhone models, no significant differences were observed in grayscale intensity distributions (ANOVA p = 0.46; Supplementary Figure 2) demonstrating cross-device robustness. Collectively, these findings support the stability and reproducibility of the standardized imaging configuration for subsequent quantitative feature extraction and ML analysis.

### On-Device Image Processing and Feature Extraction

All predefined quantitative strip features were successfully extracted on-device using native iOS image processing libraries, with processing times ranging from 42 to 58 milliseconds per image. Reproducibility was evaluated by repeated imaging of the same strip under fixed acquisition conditions. Extracted features demonstrated high stability, with intra-strip CV ranging from 3.2% to 4.8% (Table 2), indicating minimal variability attributable to optical acquisition or computational processing. Univariate analysis revealed strong discrimination between HPV-positive and HPV-negative samples across multiple quantitative features (Supplementary Table S2). Contrast-to-noise ratio (CNR) exhibited the most pronounced separation, with significantly higher values in HPV-positive samples (6.82 ± 1.45) compared with HPV-negative controls (2.14 ± 0.98; p < 0.001), corresponding to a large effect size (Cohen’s *d* = 3.88). Similarly, edge sharpness (1.92 ± 0.34 vs. 1.21 ± 0.29; p = 0.006; *d* = 2.27) and directional variance (0.87 ± 0.21 vs. 0.54 ± 0.18; p = 0.004; *d* = 1.72) demonstrated substantial effect sizes, reflecting sharper and more homogeneous signal morphology in true positive bands. Mean band intensity and test/control intensity ratio were also significantly elevated in HPV-positive samples (both p < 0.001), with large effect sizes (*d* > 1.5). These features differed significantly between HPV-positive and HPV-negative groups. However, faint-band samples near the detection limit exhibited feature values that partially overlapped with the HPV-negative distribution in univariate analyses, suggesting that single-feature thresholds may be insufficient for reliable discrimination and motivating the use of multivariable modeling.

**Table 2.** Reproducibility of Strip Features Under Repeated Imaging Conditions.

| Feature | Intra-Strip CV (%) * |
| --- | --- |
| Mean intensity | 3.2% |
| Contrast-to-noise ratio (CNR) | 4.8% |
| Directional variance | 3.9% |
| Edge sharpness | 4.1% |
| Test/control intensity ratio | 3.5% |

\* Values represent coefficient of variation (CV) calculated from repeated imaging (n = 20) of the same lateral flow strip under standardized acquisition conditions.

### ML Model Development

An initial set of 18 strip-specific quantitative features (Supplementary Table S3) was evaluated for model development. Correlation analysis identified clusters of highly collinear variables (Pearson r ≥ 0.85), as illustrated in Figure 3. To reduce multicollinearity and retain complementary predictors, seven redundant features (Median_T, SD_T, Sum_T, Min_T, Max_T, SBR, and Log_TCR) were excluded through hierarchical correlation filtering. The remaining features were subsequently subjected to LASSO regression with repeated cross-validation to identify a parsimonious predictor set. This procedure consistently selected four independent features—CNR, directional variance, edge sharpness, and the test-to-control intensity ratio—for inclusion in the final multivariable logistic regression model.

**Figure 3.**
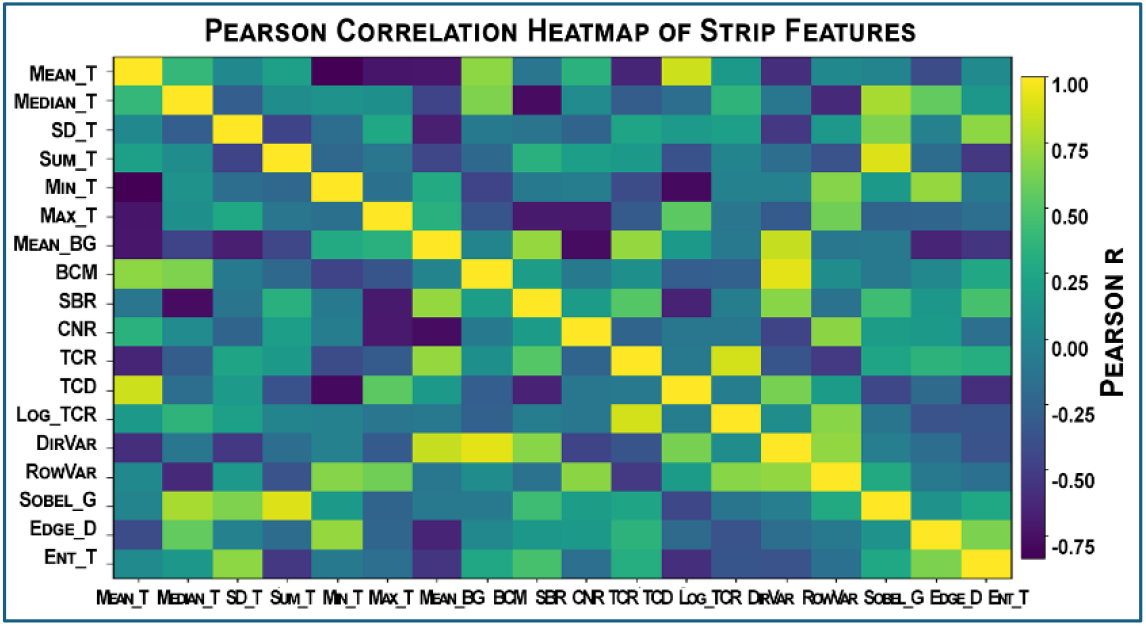
Correlation heatmap of strip-specific quantitative features. Pairwise Pearson correlation coefficients (r) were calculated among all 18 predefined strip-specific quantitative features using the development cohort. The heatmap displays the linear correlation structure among these features. Features were reordered using hierarchical clustering with complete linkage to identify correlated groups. Feature pairs demonstrating high collinearity (r≥ 0.85) were considered redundant and excluded prior to penalized regression modeling to reduce multicollinearity and enhance model stability.

A multivariable logistic regression classifier incorporating these predictors was constructed to estimate the probability of HPV positivity. The optimal classification threshold was determined using the Youden index in the development cohort and corresponded to a probability cutoff of 0.58, which maximized the combined sensitivity and specificity. This prespecified threshold was subsequently applied unchanged to the independent validation cohort to ensure unbiased evaluation of model performance. The full model equation and supporting threshold data are provided in the Supplementary Materials. As summarized in Supplementary Tables S4 and S5, the regression coefficients and feature standardization parameters enable reproducible model reconstruction and on-device inference. Model stability was further evaluated using bootstrap resampling. Across 1,000 bootstrap iterations, feature selection stability reached 92%, indicating robust retention of the selected predictors and minimal susceptibility to sampling variability.

### Model Benchmarking and Robustness Analyses

In the development cohort, the intensity-only model achieved an AUC of 0.91, whereas the multifeature model achieved an AUC of 0.986, indicating significantly improved discrimination (DeLong test, p < 0.05) (Supplementary Table S6). Sensitivity and specificity were also lower for the intensity-only model (89% and 90%) compared with the multifeature model (96.7% and 100%). These findings demonstrate that multidimensional feature integration provides substantial incremental discriminative value beyond simple band intensity quantification. To further evaluate performance relative to deep learning approaches, a lightweight convolutional neural network (CNN) was trained using identical repeated cross-validation procedures. Input images were resized to 128 × 128 pixels prior to training. The architecture comprised three 3 × 3 convolutional layers with ReLU activation, each followed by max-pooling, and a single fully connected classification layer. Optimization was performed using the Adam optimizer with binary cross-entropy loss. The CNN achieved an AUC of 0.981 in the development cohort, comparable to the multifeature logistic regression model (AUC = 0.986), with no statistically significant difference between models (p > 0.05) (Supplementary Table S7). However, CNN required more trainable parameters and lacked explicit feature-level interpretability. Thus, the interpretable logistic regression model achieves comparable performance with substantially greater transparency and efficiency.

To evaluate model performance under low-abundance signal conditions, a focused sub-analysis was performed on samples with visually ambiguous or faint CRISPR-LFA test bands. As described above, eight clinically confirmed HPV-positive samples (five HPV16 and three HPV18) contained low viral DNA levels by PCR and produced weak LFA signals that were visually interpreted as negative. The multifeature logistic regression model correctly classified six of these eight samples as HPV-positive, with predicted probabilities ranging from 0.72 to 0.86 (Supplementary Table S8). In contrast, the intensity-only model misclassified four samples as negative (predicted probabilities 0.41–0.49) and assigned near-threshold probabilities (0.55–0.63) to the remaining four cases, indicating reduced classification confidence under borderline signal conditions (Supplementary Table S8). These findings demonstrate the superior stability and discriminative ability of the multifeature model in low-signal scenarios where visual interpretation or intensity-only assessment may yield false-negative results.

### On-Device ML Inference Implementation

The final logistic regression model was embedded directly within the iOS application to enable fully self-contained, on-device inference. Prediction latency was minimal, ranging from 8 to 12 milliseconds per test. Model outputs generated on the device were numerically identical to those produced by the reference Python implementation (absolute difference < 0.001), confirming computational fidelity across platforms. Battery consumption associated with the complete workflow, including image capture, feature extraction, inference, and data encryption, was low, accounting for less than 1.8% usage per 100 tests.

### Clinical Validation of Model Discrimination and Calibration

The multivariable logistic regression model incorporating the four selected features demonstrated excellent discrimination, achieving a mean AUC of 0.986 under repeated 10×10 cross-validation, with sensitivity of 96.7% and specificity of 100% (Fig. 4; Table 3). Compared with each individual feature, the multivariable model showed significantly greater discriminative performance (Hanley–McNeil approximation, all p < 0.05; Supplementary Table S9). Bootstrap resampling yielded an optimism-corrected AUC of 0.982 (95% CI 0.973–0.991), indicating minimal overfitting and strong internal validity. In comparison, visual interpretation of CRISPR-LFA detected 67 of 75 HPV-positive cases, with eight faint bands near the detection threshold, yielding a sensitivity of 89.3% (Fig. 2; Table 3). Among these visually ambiguous cases, the logistic regression model correctly classified six (four HPV16 and two HPV18) as positive, increasing overall sensitivity to 96.7% under borderline signal conditions (Table 1).

**Table 3.**
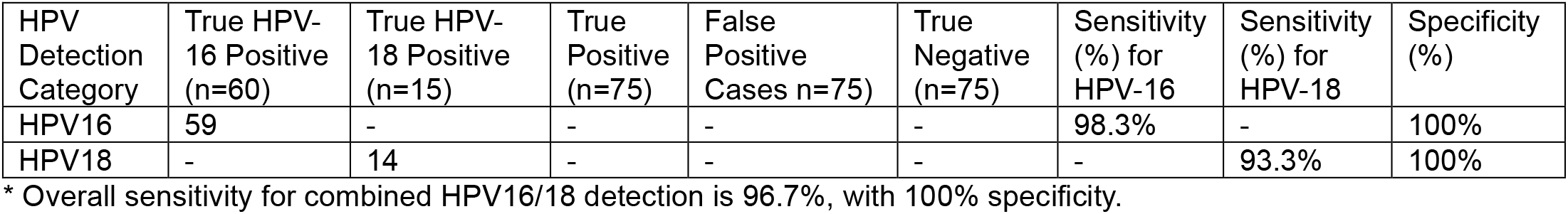
Diagnostic Performance of the Logistic Regression Model for HPV16 and HPV18 Detection in the Development Cohort ^*^.

**Figure 4.**
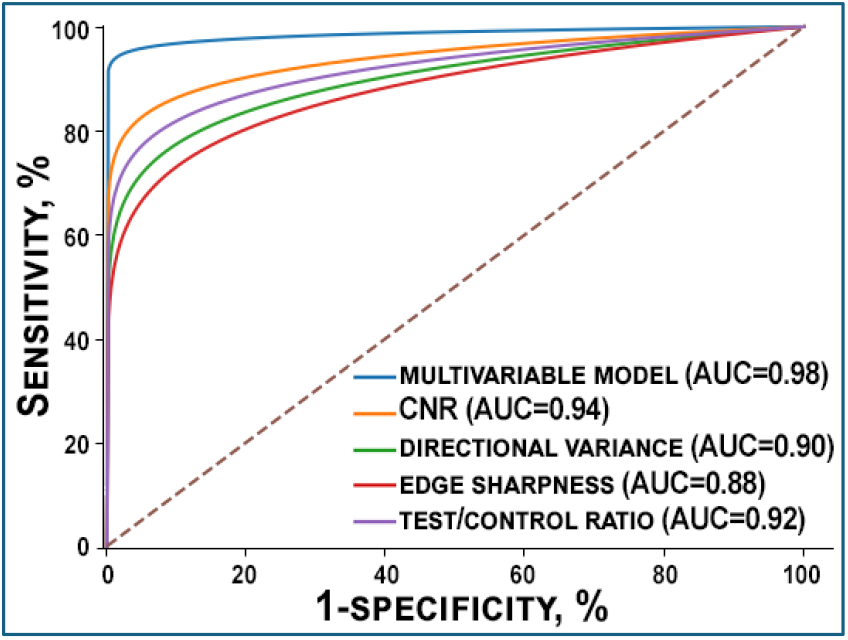
ROC curves of the multivariable logistic regression model and individual single-feature predictors (CNR, directional variance, edge sharpness, and test-to-control ratio) in the development cohort. The multivariable model demonstrated superior discriminative performance compared with each individual feature, achieving a mean AUC of 0.986 under repeated 10×10 cross-validation. Sensitivity and specificity were 96.7% and 100%, respectively. The diagonal dashed line represents chance performance (AUC = 0.5).

The model also demonstrated strong probabilistic calibration (Supplementary Figure S3), with a calibration slope of 0.97 and intercept of 0.02, indicating close agreement between predicted probabilities and observed outcomes. The Brier score was 0.028, and the Hosmer–Lemeshow test was not significant (p = 0.53), supporting good model fit (Supplementary Table 10). HPV-positive cases spanned cervical cancer stages ranging from carcinoma in situ (stage 0) to stage IV. The logistic regression model detected circulating HPV DNA across this spectrum, supporting applicability to both early- and advanced-stage disease. No significant association between cancer stage and model-predicted probability was observed (Kruskal–Wallis test, p > 0.05), indicating consistent classification performance across disease stages and potential utility for early detection (Supplementary Figure S4).

### Independent Cohort Validation and Field Deployment Robustness

Model performance was evaluated in an independent cohort of 60 plasma samples using the prespecified classification threshold of 0.58. The multivariable logistic regression model achieved an area under the AUC of 0.979 (95% CI 0.954-0.999; Supplementary Figure S5), with sensitivity of 96.7% and specificity of 100% (Supplementary Table S11). These results were consistent with internal cross-validation findings, supporting stable discrimination and preserved predictive performance in independent data. Operational robustness was further assessed across three smartphone models, three lighting stress conditions (500-900 lux), and four independent operators. Discriminative performance remained consistently high across all tested conditions, with subgroup AUC values exceeding 0.95 for each device and lighting scenario (Supplementary Table S12). Mixed-effects modeling, performed with classification accuracy as the dependent variable, demonstrated no significant device effect (p = 0.38), operator effect (p = 0.44), or device-by-lighting interaction (p = 0.29), indicating stable performance across heterogeneous acquisition environments (Supplementary Table S12). During continuous operation testing at ambient temperatures (25°C) up to 32°C, no evidence of device overheating, performance degradation, or CPU throttling was observed over a 60-minute period. Under simulated field conditions, secure local encryption and storage were maintained using native iOS data protection frameworks. In low-bandwidth environments approximating 3G connectivity, synchronization achieved a 100% upload success rate upon reconnection, with a mean transmission time of 2.3 seconds per record and no observed data corruption. Geolocation metadata was successfully captured in 96% of tests, with remaining cases attributable to user-disabled location services (Supplementary Table 13). On-device inference latency (8-12 ms), negligible numerical deviation relative to the validated Python reference implementation (<0.001), and minimal battery usage (<1.8% per 100 tests) further support computational efficiency and deployment feasibility. Consequently, these findings demonstrate preserved external generalizability, stable performance across devices, lighting conditions, and operators, and feasibility of decentralized deployment pending further field validation.

## Discussion

In this study, we developed and validated a smartphone-based CRISPR-LFA platform for sensitively and reliably detecting circulating HPV DNA. The system demonstrated excellent discrimination, robust probabilistic calibration, and consistent performance across devices, operators, and lighting conditions, while enabling fully self-contained, on-device inference. By eliminating manual band reading and generating calibrated probabilistic outputs, the system improves detection of low-intensity positives, enhances reproducibility, and supports scalable decentralized screening.

CRISPR-LFA platforms offer several advantages for detecting molecular changes, including high sensitivity, rapid turnaround, isothermal amplification, and simple lateral flow readouts that enable decentralized and POC testing^6, 11^. However, the clinical implementation remains challenging because visual interpretation of test bands is subjective and operator dependent. Weak signals near the detection limit can therefore lead to inconsistent interpretation and reduced diagnostic sensitivity. In the present study, visual interpretation identified eight samples with faint CRISPR-LFA bands near the detection threshold, resulting in false-negative classifications and reducing the overall detection sensitivity to 89.3%. In contrast, the multifeature logistic regression model correctly classified six of these visually ambiguous cases as positive, demonstrating improved sensitivity (96.7%) under low-signal conditions. Compared with conventional visual interpretation of CRISPR-Cas12a assays, this platform enables standardized quantitative analysis through objective band signal measurement and interpretable ML-based classification, thereby improving diagnostic reliability. This structured feature integration may be particularly advantageous in low–circulating tumor DNA settings, where subtle signal differences are clinically meaningful. Therefore, our present findings are clinically relevant because circulating HPV DNA detected in plasma may complement cervical swab–based HPV testing by serving as a liquid biopsy biomarker that reflects tumor burden or disease progression. Furthermore, the smartphone-enabled CRISPR platform could facilitate decentralized testing for patient triage or disease monitoring in POC settings with limited laboratory infrastructure.

Recent studies have applied CNN-based approaches to automate CRISPR-LFA interpretation, primarily in infectious disease settings ^11, 12, 19^. Although this approach can achieve high accuracy, it often requires large training datasets and complex models that are difficult to interpret and calibrate, which may complicate clinical validation and regulatory approval for POC use. Given our moderate sample size and emphasis on transparent probabilistic outputs, we adopted a radiomics-inspired feature engineering strategy^17^. Interpretable quantitative descriptors—including contrast-to-noise ratio, directional variance, edge sharpness, and the test-to-control intensity ratio—were extracted from standardized strip regions. These features were then integrated into a parsimonious multivariable logistic regression model after structured feature reduction and internal validation. Although individual features showed strong univariate discrimination, their multivariable integration substantially improved model performance, yielding an AUC of 0.986 in both the development and independent validation cohorts. Furthermore, compared with a lightweight CNN, the structured radiomic feature–based framework provides greater interpretability, calibration transparency, and computational efficiency, offering a practical alternative to more complex deep learning models.

Another key implementation feature of this system is fully on-device inference without reliance on proprietary software development kits, cloud computation, or third-party AI services. Model coefficients and preprocessing parameters were embedded directly within the smartphone application, ensuring deterministic predictions with minimal latency (<12 ms) and computational fidelity relative to the validated Python reference implementation. Stable performance was confirmed in an independent external validation cohort and across varied acquisition conditions, with no significant device or operator effects observed. Furthermore, the prototype hardware costs were approximately $22 USD and are projected to decrease below $15 USD with scaled manufacturing, supporting feasibility for scalable public health deployment. In addition, although this study used iPhone devices, the framework can be adapted to other smartphones with appropriate calibration. Moreover, compared with conventional HPV detection methods (Supplementary Table 14), the CRISPR-ML platform may provide high diagnostic sensitivity, faster turnaround time (∼40 min), and minimal equipment requirements through smartphone-based operation, supporting decentralized POC testing.

Several limitations should be considered. Although the developed platform shows promise, larger multicenter studies involving diverse and asymptomatic populations will be necessary to validate its performance in broader screening settings. Furthermore, the current implementation relied on a light-controlled enclosure to standardize image acquisition; therefore, performance under fully uncontrolled field conditions remains to be evaluated. In addition, validation of the enclosure in real-world environments and across additional devices will be necessary to confirm operational robustness and monitor potential calibration drift. Moreover, future studies may further expand this framework through longitudinal surveillance modeling, integration with electronic health systems, incorporation of additional biomarkers, and exploration of hybrid feature–deep learning approaches.

## Conclusion

This easily interpretable and smartphone-based CRISPR-LFA platform demonstrates high diagnostic accuracy, robust calibration, and stable performance across diverse acquisition conditions. The platform shows promise for scalable cervical cancer screening and early detection in POC settings and has broad potential for extension to other disease applications.

## Supporting Information

The Supporting Information is available free of charge on the ACS Publications website. Supplementary Figures S1–S5, Supplementary Tables S1–S14, and additional experimental details (PDF).

## Ethics approval and consent to participate

The study protocol was reviewed and approved by the Advarra Institutional Review Board (Advarra, Columbia, MD, USA; protocol number 22-8356). Peripheral blood samples were collected from human participants following written informed consent and processed to obtain plasma for analysis. All procedures involving human participants were conducted in accordance with the ethical standards of the institutional research committee and with the Declaration of Helsinki and its later amendments. Plasma samples were anonymized prior to analysis to protect participant confidentiality.

## Consent for publication

Not applicable.

## Availability of data and materials

The datasets generated and/or analyzed during the current study are available from the corresponding author upon reasonable request.

## RediT authorship contribution statement

Jipei Liao: Data curation, Investigation, Methodology.

Jesse Rima: Data curation, Formal analysis, Investigation, Methodology, Validation, Visualization.

Aarush Sharma: Software, Methodology, Formal analysis, Data curation. Jen-Hui Tsou: Methodology, Validation, Supervision.

Feng Jiang: Conceptualization, Resources, Supervision, Project administration, Writing –review & editing.

## Acknowledgments

The authors thank the participants who contributed plasma samples for this study.

## Appendix A

**Supplementary data**

Supplementary data associated with this article can be found in the online version.

## Data availability

All data generated or analyzed during this study are included in this article (and its supplementary information files).

